# Development and validation of a clinical prediction model of domain-specific post-stroke cognitive impairment

**DOI:** 10.1101/2024.09.06.24313196

**Authors:** Andrea Kusec, Kym IE Snell, Nele Demeyere

**Affiliations:** Nuffield Department of Clinical Neurosciences, University of Oxford, Oxford, UK; Department of Applied Health Sciences, School of Health Sciences, University of Birmingham, Birmingham, UK; National Institute for Health and Care Research (NIHR) Birmingham Biomedical Research Centre, UK

**Keywords:** stroke, cognitive dysfunction, cognitive impairment, prediction modelling, risk prediction

## Abstract

**Background and Objectives:** Post-stroke cognitive impairment (PSCI) is highly prevalent across multiple cognitive domains. Individualised PSCI prognosis has mainly been researched using outcomes not developed specifically for stroke (i.e., dementia risk). Further, existing models often use predictors not routinely available in electronic health records. Here, we develop and externally validate clinical prediction models for overall PSCI using a stroke-specific cognitive outcome using acute cognition and data routinely collected as part of stroke care.

**Methods:** *N*=430 stroke survivors completed the Oxford Cognitive Screen (OCS) in acute care and at 6-month follow-up (binarized outcome; impaired vs unimpaired). Logistic regression models were fitted comprising both mandatory clinically-relevant (age, sex, stroke severity, education, stroke hemisphere, acute PSCI) and data-driven (acute mood difficulties, length of stay in acute care, multimorbidity) predictors using backward elimination (*p* < 0.10) on multiply imputed data. Internal validation used bootstrapping to obtain optimism-adjusted performance estimates. External validation used the optimism-adjusted C-Slope as a uniform shrinkage factor.

**Results:** The overall PSCI model demonstrated good optimism-adjusted performance (C-Statistic=0.76 [95% CI=0.71–0.80]). In external validation, the overall PSCI model was comparable to development data (C-Statistic=0.74 [95% CI=0.67–0.79]). PSCI model performance did not vary by sex, but performed best in adults <60 years old (C-Statistic=0.76) with moderate-severe acute PSCI (C-Statistic=0.72). We further explored modelling of

**Discussion:** Prediction models of stroke-specific cognition have the potential to offer more meaningful PSCI prognoses, including domain-specific cognitive recovery, compared to existing models focused on domain-general decline. The present model shows promise of generalisability with an initial good external validation performance of the PSCI model in a different stroke severity cohort. Future recalibration of domain-specific models would be beneficial to provide additional detailed predictions.

## Introduction

Stroke is the leading cause of long-term physical and cognitive disability worldwide^1^. One-year post-stroke cognitive impairment (PSCI) prevalence estimates range from 40%^2^ to 98%^3^, with rates of 45%^4^ to 80%^5^ in chronic stroke. PSCI negatively impacts patients^6^, caregivers/families^7^, and has considerable economic costs^8^.

Clinical prediction models have been developed to improve PSCI prognostication, chiefly post-stroke cognitive decline and dementia^9,10^. However, PSCI does not necessarily cause cognitive decline or dementia. Research demonstrates that whilst some patients exhibit decline, others have a stable, chronic cognitive impairment or even demonstrate continued improvement^11^. New PSCI definitions acknowledge the complex interplay of declining brain health, focal brain injury and cognitive recovery, with outcomes including decline, stability, and improvement^12^. PSCI is highly prevalent across multiple cognitive domains of language, memory, attention, numeracy, executive function, and praxis^13^. These impairments have previously been studied in isolation, despite research demonstrating differing recovery rates across domains (e.g., hemispatial neglect^14^).

Existing prediction models of post-stroke dementia^15–17^ perform poorly in PSCI^18,19^, possibly because specific cognitive domains have different relationships to functional outcomes^20^. Newer models have attempted to improve PSCI prediction, but remain focused on traditional clinical and demographic predictors (e.g., age, stroke severity). A recent meta-analysis (*N*=160,783) of PSCI predictors reported that, by far, the strongest predictor of chronic PSCI was acute cognitive functioning^21^, demonstrating the importance of baseline cognitive performance in developing accurate prediction models^22^. However, existing PSCI prediction models do not routinely include acute cognition as a predictor. With early PSCI assessment now recommended by national and international guidelines^23–25^ acute cognitive data should be routinely available and used in PSCI prognostication.

Existing PSCI prediction models often assess acute and long-term cognition via dementia screening tools (e.g., Mini Mental State Examination). These tools are not suitable for left hemisphere stroke due to overreliance on verbal abilities^26^. Utilizing a stroke-specific cognitive screen for PSCI clinical prediction models would avoid assessment confounds and is more strongly associated with 6-month cognitive recovery^13,27^.

## Study Aims

To develop and externally validate clinical prediction models of 6-month PSCI outcomes using acute cognitive information via a stroke-specific tool.

## Methods

The study is a secondary analysis of data collected from the Oxford Cognitive Screening Programme. All participants provided informed consent to take part (REC Reference: 18/SC/0550).

### Participants

Participants comprised a single, consecutively recruited cohort from the John Radcliffe Hospital acute stroke ward between March 2012 (first consented participant) to March 2020 (final follow-up participant).

Programme inclusion criteria were 1) stroke diagnosis (first ever or recurrent; determined via CT/MRI scan); 2) ≥18 years; 3) ability to remain alert for ≥20 minutes, and 4) ability to provide informed consent. Participants completed a brief stroke-specific cognitive assessment acutely (*N=*866) and at 6-months post-stroke (*N=*430). Attrition from acute to 6-months was due to loss to follow-up (*n*=149), participant death (*n*=82), declining follow up (*n=*76), being too unwell to take part (*n*=72), moved out of area (*n*=36), had an incomplete assessment (*n=*16), or could not be seen due to COVID-19 restrictions (*n*=5). Those lost to attrition versus those retained only differed in terms of having greater stroke severity and greater acute cognitive impairments. A detailed account of attrition is in Milosevich et al.^13^

Stroke severity (National Institute of Health Stroke Severity; NIHSS) and other stroke-related details (e.g., lesion hemisphere, first vs recurrent stroke) were obtained from electronic health records.

### Study Outcome Measure

Overall PSCI was assessed using the ***Oxford Cognitive Screen*** (OCS^28^), a measure of stroke-specific cognition across multiple cognitive domains affected by stroke. The OCS was administered at a mean of 4.38 days (*SD* = 4.46) post-stroke

The OCS comprises 12 subtasks forming 6 cognitive domains: Language (picture naming, semantics, sentence reading), Memory (orientation, verbal recall, episodic recognition), Spatial attention (broken hearts cancellation task), Numeracy (number writing, calculation), Praxis (gesture imitation), and Executive Function (mixed trails). Subtask scores are binarized as impaired (1) or unimpaired (0) relative to cut-off scores from a normative sample. A domain impairment was defined as the presence of any impairment in any subtask within that domain.

Models developed included overall PSCI severity (total proportion of OCS subtasks impaired at 6-months post-stroke; continuous outcome model), and binarized PSCI presence in any domain (logistic outcome model).

### Analysis

Analyses were performed in R Version 4.4.0^29^. Baseline descriptive statistics were first summarised. R packages used included *rms*^30^, *psfmi*^31^, *mice*^32^, and *pmvalsampsize*^33^. Data is freely available at https://www.dementiasplatform.uk/ and analysis code at https://osf.io/3pc5k/

### Predictor Selection

We selected predictors likely to be available in electronic health records upon deployment. This included clinically relevant predictors (age at stroke, sex, NIHSS scores, education years, first vs recurrent stroke, type of stroke [ischaemic vs haemorrhagic], stroke hemisphere, and acute cognitive impairment^21^). The predictor stroke hemisphere included an “undetermined” category where either lateralisation was not clear based on CT/MRI scan data and/or clinical presentation was not conclusive. We additionally included “data-driven predictors” that are available in electronic health records (length of stay in acute care, independence prior to admission [defined as not requiring paid or family carer support >2 hours per week], presence of mood difficulties during acute care [as reported by clinical records during stay], and Charlson Comorbidity index). Each model therefore had 13 potential predictors forming an “initial model.” Following predictor selection, performance was estimated with clinically relevant predictors and only significant data-driven predictors retained, labelled throughout the manuscript as the “final model.” Predictor selection per model was repeated using bootstrapping across 1000 iterations.

### Sample Size Justification

Sample size sufficiency was evaluated based on the events fraction, total sample size, number of predictor parameters, and a target shrinkage factor of >0.90 to minimise overfitting^34,35^.

Event fraction rates and assumed apparent Nagelkerke’s R^2^ performance (0.30) were based on previous Oxford Cognitive Screening programme analyses^13^. For the overall PSCI model (*N* Events=295 of 430), the sample size required was 393 participants, indicating sufficient power with the current data.

### Missing Data Management

Those with complete vs incomplete data at 6-months post-stroke were compared on predictor variables. To increase statistical power and reduce bias, multiple imputation was conducted across 20 imputed datasets (due to 28.8% missingness in acute NIHSS scores) with 50 iterations. Data were assumed missing at random given that the variable with the highest missingness rate (NIHSS) was historically unavailable in electronic health records in earlier recruitment periods. Only predictor variables were imputed^36^. Upon model deployment, missingness is likely to occur (e.g., missing stroke severity information) and therefore imputation would be necessary^36^. Sensitivity analyses were conducted investigating the influence of missing information. A detailed account of participant attrition in this cohort is elsewhere^13^.

### Model Development and Internal Validation

Across all models, clinically relevant predictors were retained irrespective of statistical significance. Backward stepwise elimination was used to remove only non-significant (*p*>0.10) data-driven predictors. This approach was taken given criticisms around removing clinically relevant (though statistically insignificant) predictors^37^. Predictor selection was conducted only on data-driven predictors, as clinically relevant predictors were viewed to be mandatory to retain due previous research demonstrating associations to 6-month PSCI, their conceptual importance, and face validity for clinicians using the model upon deployment. Models developed across 20 imputed datasets were compared to complete case data.

Apparent final model performance (i.e., non-significant data-driven predictors removed) was evaluated using discrimination (model’s ability to correctly identify individuals with and without 6-month PSCI; estimated via the Area Under the Curve [AUC; binary outcome models only], C-Statistic), calibration measures (calibration-in-the-large [CITL], calibration slope [C-Slope], calibration plots, Brier scores, observed-expected ratio) and goodness-of-fit measures (adjusted R^2^, continuous PSCI model; Nagelkerke’s R^2^, binary PSCI models). Pooled (across imputed datasets) b-values, odds ratios (ORs), and performance statistics are reported per model.

Optimism-adjusted performance estimates were obtained via bootstrapping each model on multiply imputed data across 1000 iterations. The model-specific optimism-adjusted C-Slope was used as a uniform shrinkage factor and was multiplied with model regression coefficients to correct for potential overfitting^38,39^. Model intercepts were re-estimated using the shrunken regression coefficients to obtain an accurate CITL.

Risk groups were created using 10^th^ decile groups on prediction model estimates for visualisation purposes via calibration plots.

### External Validation

The OCS-Care dataset^11^ was used for external validation. In parallel to OCS-Recovery data, the OCS-Care dataset (*N*=264, *M* age = 68.9) assessed acute PSCI using the OCS and 6-months later, comprising a mild severity cohort (*Mean* NIHSS=2.8). Model predictors were collected from electronic health records. Many OCS-Care participants had at least one cognitive impairment at 6-months (*N* events=147; 55.6%), versus 70.8% acutely (*N* events=187). When estimating minimum sample size requirements^40^, this dataset was sufficient to estimate a C-Statistic of 0.80 (CI width=0.20), though precise C-Slope estimates require a much larger dataset.

Shrunken model coefficients obtained through internal validation were applied to the OCS-Care data to estimate performance. Performance measures described above (C-Slope, CITL, C-Statistic, R^2^ and Nagelkerke’s R^2^) were estimated. Overall binary PSCI model performance was further evaluated within subgroups by age range, sex, first vs recurrent stroke, and acute PSCI severity.

### Exploratory Domain-Specific Prediction Models

Because the OCS provides information on domain-specific cognition on language, attention, memory, numeracy, praxis, and executive function, we additionally explored creating logistic domain-specific prediction models using the same development and validation methods detailed above for the overall PSCI model as a signal for future development of domain-specific models. Given the sample size requirements were not met for individual subdomains of Language (1000 participants; *N* Events=138 of 428), Memory (1016 participants; *N* Events=137 of 430), Attention (638 participants; *N* Events=187 of 416), Praxis (2912 participants; *N* Events=79 of 403), Number (2524 participants; *N* Events=85 of 420), and Executive Function (1724 participants; *N* Events=98 of 400) models, these results are presented in the *Supplementary Materials*.

## Results

Demographics are in Table 1. At 6-months, all participants provided outcome data.

**Table 1.** Participant demographics. NIHSS=National Institute of Health Stroke Severity

| Participants (N=430) |  | Min-Max |
| --- | --- | --- |
| **Sex**—$n$ (%) | | |
| Male | 230 (53.5%) |  |
| Female | 200 (46.5%) |  |
| **Age**—Mean (SD) | 73.8 (12.5) | 18–95 |
| **Education Years**—Mean (SD) | 12.3 (3.6) | 0–23 |
| **Stroke Type**—$n$ (%) | | |
| Ischaemic | 362 (84.2%) |  |
| Haemorrhagic | 68 (15.8%) |  |
| **Lesion Hemisphere**—$n$ (%) | | |
| Left | 153 (35.6%) |  |
| Right | 168 (39.1%) |  |
| Bilateral | 34 (7.9%) |  |
| Undetermined from scan | 75 (17.4%) |  |
| **First or Recurrent Stroke**—$n$ (%) | | |
| First | 292 (67.9%) |  |
| Recurrent | 138 (32.1%) |  |
| **Acute NIHSS**—Mean (SD) | 6.8 (6.1) | 0–30 |

Participants with missing 6-month PSCI data were more likely to have higher acute PSCI (*p*<0.0001), be older in age (*p*=0.02), and have acute language (*p*<0.01), memory (*p*<0.001), or numeracy impairments (*p*<0.001). NIHSS missingness was not related to demographic factors (*p*s>0.15; see *Supplemental Materials*).

### Overall PSCI Models

Pooled shrunken coefficients of the final overall continuous and binary PSCI models are in Table 2.

**Table 2.** Pooled model coefficients of continuous and binary overall PSCI models. Shrinkage was applied to coefficients using the optimism-adjusted C-Slope. For categorical variables stroke hemisphere and independence pre-admission, reference categories were left-hemisphere stroke and independent pre-admission, respectively. NIHSS=National Institute of Health Stroke Severity; OCS=Oxford Cognitive Screen

|  | Overall Score (severity-continuous) |  | Any cognitive impairment (binary) |  |
| --- | --- | --- | --- | --- |
|  | B [95% CI] | p-value | OR [95% CI] | p-value |
| **Clinically Relevant Predictors** |  |  |  |  |
| **Intercept** | -0.05 [-0.15–0.06] | 0.33 | 0.12 [0.02–0.72] | <0.05 |
| **Stroke Age** | 0.002 [0.001–0.003] | <0.01 | 0.93 [0.92–0.93] | <0.001 |
| **Sex (Female)** | 0.01 [-0.01–0.04] | 0.38 | 0.94 [0.87–1.03] | 0.57 |
| **Education Years** | -0.003 [-0.01–0.001] | 0.19 | 0.91 [0.89–0.92] | 0.08 |
| **NIHSS** | 0.001 [-0.002–0.003] | 0.55 | 0.92 [0.91–0.93] | 0.59 |
| **Hemisphere** |  |  |  |  |
| Right | 0.01 [-0.02–0.04] | 0.63 | 0.93 [0.84–1.02] | 0.89 |
| Bilateral | -0.05 [-0.10–0.001] | 0.06 | 0.75 [0.64–0.88] | <0.05 |
| Undetermined from Scan | 0.01 [-0.03–0.05] | 0.73 | 0.86 [0.76–0.97] | 0.32 |
| **First/Recurrent Stroke** |  |  |  |  |
| Recurrent Stroke | 0.02 [-0.01–0.05] | 0.14 | 1.01 [0.92–1.10] | <0.05 |
| **Stroke Type** |  |  |  |  |
| Haemorrhagic | -0.02 [-0.06–0.01] | 0.22 | 0.89 [0.80–1.01] | 0.67 |
| **Acute Proportion OCS Tasks Impaired** | 0.35 [0.29–0.41] | <0.001 | 1.62 [1.35–1.95] | <0.001 |
| **Data Driven Predictors** |  |  |  |  |
| **Independence Pre-Admission** |  |  |  |  |
| Carer Support | 0.10 [0.05–0.14] | <0.001 | -- | -- |
| Family Support | 0.04 [-0.03–0.10] | 0.25 | -- | -- |

In the multivariable continuous model of proportion of 6-months OCS tasks impaired, the strongest clinically relevant predictors included higher age (pooled *B*=0.002 [95% CI=0.001– 0.003]) and a greater proportion of acute OCS tasks impaired (pooled *B*=0.35 [95% CI=0.29– 0.41]). In bootstrapped and complete case data, the only data-driven predictor retained was requiring carer support prior to admission (pooled *B*=0.10 [95% CI=0.05–0.14]) and improved model fit in complete case data (*F*=3.69, *p*=0.03). The optimism-adjusted performance of the continuous overall PSCI model was good to excellent (C-Slope=0.96, CITL=-0.01; MSE=0.02; Adj R^2^=0.34).

In the multivariable binary model, higher age (pooled OR=0.93 [95% CI=0.92–0.93]), bilateral hemisphere lesions (pooled OR=0.75 [95% CI=0.64–0.88]), fewer years of education (pooled OR=0.91 [95% CI=0.89–0.92]), and a greater proportion of acute OCS tasks impaired (pooled OR=1.62 [95% CI=1.35–1.95]) were associated with an increased risk of 6-month PSCI. No data-driven predictors were retained for the binary PSCI model. The final optimism-adjusted performance showed good performance (C-Statistic=0.76 [95% CI=0.71–0.80]; C-Slope=0.93 [95% CI=0.75–1.11]; CITL= −1.17 [95% CI= −1.39– −0.95]; Brier Score=0.12 [95% CI=0.10–0.14]; Nagelkerke’s R^2^=0.21).

In sensitivity analyses using complete case data, there were no notable differences in predictor selection for either the continuous or binary overall PSCI models.

Unadjusted relationships between predictor and outcome variables for the overall PSCI models are in *Table S11.* All model performance measures for overall PSCI are in Table 3.

**Figure 1.**
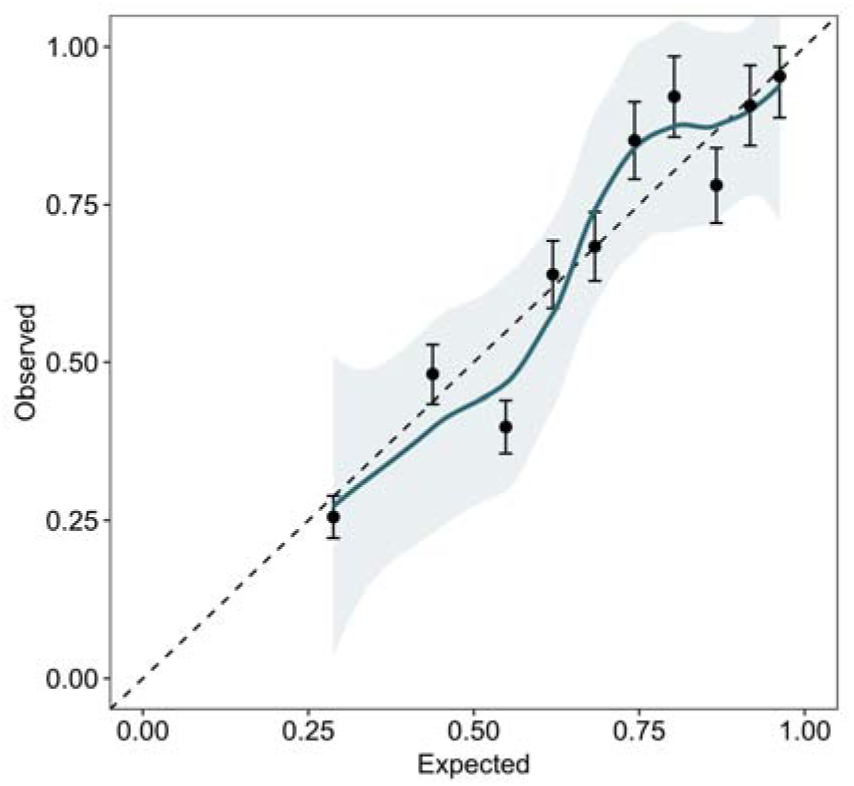
Calibration plot of any 6-month PSCI (0=no impairment, 1=any impairment) across 20 imputed datasets. Complete cases plot is in the *Supplementary Figure S1*.

**Table 3.** Performance metrics all final models pooled across 20 imputed datasets. CITL=Calibration-in-the-large; E/O=Expected:Observed Ratio; C-Slope=Calibration Slope

| Model | Estimate | Model Performance [95% CI] | Average Optimism [95% CI] | Optimism-Adjusted Performance [95% CI] |
| --- | --- | --- | --- | --- |
| **Overall PSCI (Continuous)** | C-Slope | 1 [0.88–1.12] | 0.04 | 0.96 |
|  | CITL | 0.00 [-0.0001–0.0001] | -0.01 | -0.01 |
|  | MSE | 0.02 [-0.05–0.08] | 0.00 | 0.02 |
| | Adjusted $R^2$ | 0.38 [0.31–0.45] | 0.04 | 0.34 |
| **Any domain impairment (Binary)** | C-Statistic | 0.78 [0.73–0.83] | 0.02 [0.02–0.03] | 0.76 [0.71–0.80] |
|  | C-Slope | 1.03 [0.85–1.22] | 0.10 [0.09–0.11] | 0.93 [0.75–1.11] |
|  | CITL | 0.0001 [-0.04–0.04] | 1.17 [0.91–1.35] | -1.17 [-1.39– -0.95] |
|  | Brier Score | 0.17 [0.15–0.19] | 0.05 [0.04–0.05] | 0.12 [0.10–0.14] |
|  | E/O | 1 [1–1] | 0.01 [-0.06–0.06] | 0.99 [0.94–1.06] |
| | Nagelkerke's $R^2$ | 0.27 | 0.06 [0.06–0.07] | 0.21 |

### External Validation

External validation estimates are in Table 4. Model discrimination, calibration, and goodness-of-fit was not substantially discrepant in the OCS-Care dataset, suggesting good model performance across cohorts.

**Table 4.** Model performance following external validation across 20 imputed datasets using OCS-Care data. CITL=Calibration-in-the-large; C-Slope=Calibration Slope

| Model | Estimate | Model Performance [95% CI] |
| --- | --- | --- |
| **Overall PSCI (Continuous)** | C-Slope | 1.26 [1.03–1.49] |
|  | CITL | -0.08 [-0.04– -0.12] |
|  | MSE | 0.01 [0.009–0.017] |
|  | Adjusted R<sup>2</sup> | 0.46 |
| **Any domain impairment (Binary)** | C-Statistic | 0.74 [0.68–0.80] |
|  | C-Slope | 1.10 [0.73–1.48] |
|  | CITL | -0.13 [-0.42–0.15] |
|  | Nagelkerke's R<sup>2</sup> | 0.22 |

In the binary overall PSCI model subgroup analyses (Table 5), performance did not vary by sex (Male C-Statistic=0.76 [0.67–0.83], Female C-Statistic=0.76 [0.66–0.84]) or by first versus recurrent stroke (First-ever C-Statistic=0.75 [0.67-0.81]; Recurrent C-Statistic=0.73 [0.57–0.84]. Model performance varied by age group (<60 years C-Statistic=0.76 [0.62–0.86], >60 years C-Statistic=0.65 [0.48–0.78], >70 years C-Statistic=0.65 [0.49–0.78], >80 years C-Statistic=0.71 [0.52–0.72]), and by level of acute PSCI (Mild acute PSCI C-Statistic=0.62 [0.52–0.72]; Moderate-severe acute PSCI=0.72 [0.61–0.81]).

**Table 5.** Subgroup analysis of overall binary cognitive model performance in external data. PSCI = Post-stroke cognitive impairment

| Subgroup | C-Statistic<br>[95% CI] | C-Slope<br>[95% CI] | Nagelkerke's R<sup>2</sup> |
| --- | --- | --- | --- |
| Males | 0.76 [0.67 – 0.83] | 1.29 [0.72 – 1.86] | 0.24 |
| Females | 0.76 [0.66 – 0.84] | 1.27 [0.67 – 1.88] | 0.18 |
| <60 years | 0.76 [0.62 – 0.86] | 1.70 [0.72 – 2.67] | 0.28 |
| >60 years | 0.64 [0.49 – 0.77] | 1.07 [0.09 – 2.04] | 0.11 |
| >70 years | 0.65 [0.50 – 0.78] | 0.64 [-0.09 – 1.37] | 0.06 |
| >80 years | 0.71 [0.54 – 0.84] | 1.24 [0.15 – 2.33] | 0.15 |
| First-Ever Stroke | 0.75 [0.67 – 0.81] | 1.24 [0.77 – 1.70] | 0.24 |
| Recurrent Stroke | 0.73 [0.57 – 0.84] | 0.97 [0.14 – 1.81] | 0.13 |
| Mild acute PSCI | 0.62 [0.51 – 0.71] | 0.79 [0.15 – 1.43] | 0.03 |
| Moderate-severe acute PSCI | 0.72 [0.60 – 0.81] | 0.99 [0.38 – 1.61] | 0.14 |

We developed an online risk calculator of the binary overall PSCI model given its promising performance https://ocs-strokecogpredictor.shinyapps.io/OCS_Stroke_Cognition_Predictor/ allowing further independent evaluation in future research. The tool allows entry of raw values of each key predictor, resulting in a percentage likelihood of domain-specific PSCI impairment at 6-months post-stroke.

### Exploratory Domain-Specific Prediction Models

Optimism-adjusted performance of the exploratory domain-specific models of PSCI were either similar to overall PSCI models (Language, Attention; C-Statistic range=0.73–0.77) or had lower performance (Memory, Numeracy, Executive Function, and Praxis; C-Statistic range=0.60–0.71). In external validation, Language and Attention models demonstrated good discrimination and goodness-of-fit, however calibration was poor across all domains, particularly in Memory, Numeracy, Executive Function, and Praxis. Full details of the exploratory domain-specific models, including external validation, are shown in the *Supplementary Materials*.

## Discussion

We developed and externally validated overall PSCI prediction models, utilising acute cognitive information from a stroke-specific screen (OCS) alongside established PSCI predictors. To our knowledge, this is the first study to use acute cognitive data and a stroke-specific measure of PSCI in clinical prediction model development. We additionally explored creating domain-specific prediction models to be developed in future research.

### Overall Model Performance

Our models provided good explanatory power, with optimism-adjusted C-Slopes of 0.95 and 0.76 for continuous and binary overall PSCI models, respectively. Compared to models of post-stroke decline and dementia (C-Statistic range=0.53–0.66^19^), our models comparatively perform better even when considering domain-specific models (C-Statistic range=0.60–0.77). Promisingly, in external data, overall PSCI model performance was comparable to internal validation, suggesting it could be used across different stroke cohorts. The domain-specific PSCI model performance showed some variation within subgroups, with a better performance in younger age groups (<60) and with moderate-severe acute PSCI. This is in line with the nature of these domain-specific PSCI predictions being indicative of stroke-specific focal cognitive changes and their recovery, rather than overall, non-stroke specific brain health related cognitive outcomes (e.g. dementia/decline).

In external data, CITL estimates were consistently negative, suggesting a small systematic overprediction of 6-month PSCI risk. This is likely due to development data comprising a more moderate-severe stroke cohort, whilst external data comprised a more mild stroke cohort. Additionally, C-Slopes in external data were larger, potentially indicating overshrinkage. This prediction model should further be recalibrated across a range of stroke severities.

### Clinical Implementation of Overall PSCI Models

Age, sex, years of education, NIHSS scores, recurrent stroke and stroke type contribute to PSCI^21^ and should be included in recalibrations of our models. Novel predictors should also be considered; our modelling approach includes data-driven predictors to allow for routine model updating. This approach identified potential predictors for recalibration (i.e., requiring carer support pre-stroke and comorbidity) that are currently excluded from PSCI prediction modelling^9,19,41,42^. Crucially, we only selected predictors available in electronic health records, in order to facilitate future implementation. Many PSCI models often include predictors not routinely available at deployment^9,10,19^. In the UK specifically, predictor selection should be guided by the National Clinical Guideline for Stroke^25^ such as including acute cognitive assessment, as these are more likely to available. Other biopsychosocial (e.g., white matter hyperintensities, socioeconomic status) and clinical (e.g., amount/intensity of neurorehabilitation offered) predictors may improve model performance, however these may be less available or have significant economic considerations. For example, imaging-based data improves PSCI prediction models^43^, however behavioural data (e.g., cognitive assessments) is considerably more affordable and feasible to implement^44,45^.

As typical for electronic health record data, NIHSS scores had large amounts of missingness. Imputation methods should be considered at deployment^36^. Collecting feedback on model usability given predictor missingness (e.g., Archer et al.^46^) would aid future implementation.

### Role of Overall Clinical Prediction Models in PSCI Prognoses

Clinical prediction models using acute cognitive information may offer more meaningful PSCI prognoses, as evidenced by better model performance compared to published prediction models of post-stroke cognition. Though PSCI rates are highest during acute stroke, early PSCI may be reversible^47^ and information about likely 6-month outcomes is valuable to stroke patients^48^. Qualitative research suggests that focusing solely on cognitive decline as a possible PSCI outcome (e.g., Hbid et al.^9^) may cause undue concern or at best be irrelevant^49^. Stroke survivors and families commonly report wanting personalized information about managing cognitive changes that are personalised to their circumstances^48^. Our models are an essential first step to providing person-specific cognitive trajectories.

### Strengths and Limitations

A notable strength is using a stroke-specific PSCI outcome measure rather than an overall cognitive decline or dementia score. The OCS’ minimisation of confounds, brief administration time, and information on cognitive domains compromised in stroke (Language, Memory, Attention, Numeracy, Executive Function and Praxis) make it a credible candidate for PSCI model development.

Though our overall PSCI performs well and is an improvement on cognitive decline-focused models, domain-specific models may provide further personalised detail still. Low prevalence^50^ of certain domain-specific outcomes (numeracy, executive function, and praxis) restricted development of domain-specific models. Whilst highly common acutely, these impairments are less prevalent at the chronic stage and therefore require substantial sample sizes for sufficient development. Future domain-specific models could also consider how combinations of specific cognitive impairments influence performance. Specific impairment combinations (e.g., language and executive function) could however affect outcomes differently, given differential correlations between cognitive domains^13^. Developing within-domain models (e.g., sentence reading model vs language impairment model) may also be helpful, given varying recovery within domains^4,13^. Furthermore, predictor selection should be carefully considered. Baseline cognition best explains long-term PSCI risk^21^ with established predictors other than age explaining little variance^13,42^. Less frequently researched PSCI domains (e.g. numeracy, praxis) may particularly benefit from data-driven predictors.

Finally, as typical for new prediction models, recalibration of our models is required.

## Conclusion

We demonstrate that a domain-specific model including acute cognitive information improved prediction of 6-month PSCI, with initial external validation in a milder cohort. Our model development process allows for future inclusion of novel data-driven predictors. This prediction model of stroke-specific cognition has the potential to offer more meaningful PSCI prognoses, including domain-specific cognitive recovery, compared to existing models focused on domain-general decline.

## Data Availability

Data is freely availble at DementiaPlatforms UK https://dementiasplatforms.uk/

