## Supplementary Material for "Development and validation of a clinical prediction model of domain-specific post-stroke cognitive impairment"

**Supplementary Materials**

***Missingness Sensitivity Analyses***

In general, missingness was minimal in 6-month cognitive outcomes (Language *N* missing = 2 of 430; Attention missing *N =* 17 of 430; Praxis missing *N* = 27 of 430; Numeracy missing *N* = 10 of 430; Executive Function missing *N* = 30 of 430), with no missing information on 6-month memory impairments or on overall PSCI. Participants with any missing information at 6-months were on average older (Welch’s *t* = -2.27, *p =* 0.03) and had a greater severity of acute PSCI (Welch’s *t* = -4.07, *p* < 0.001) but otherwise did not differ in terms of sex (ꭓ^2^ = 2.50*, p* = 0.11), type of stroke (ꭓ^2^ = 0.34*, p* = 0.56), years of education (Welch’s *t* = 1.71, *p =* 0.09), or acute NIHSS scores (Welch’s *t* = -1.03, *p =* 0.31).

When examining missingness by cognitive domain, there was some bias observed in the attention and numeracy domains. Those with 6-month numeracy data were more likely to be older (*p <* 0.05) and were more likely to have no acute numeracy impairments (*p <* 0.01).

| **Measure** | **Complete 6-Month Language (*N* = 428)** | **Missing 6-Month Language (*N* = 2)** | **Test Statistic** |
| --- | --- | --- | --- |
| **Demographics**  Age  Sex  Stroke Type  Years of Education  Acute NIHSS Score  Acute Language Impairment | *M =* 73.87 (12.50)  53.02% Male  84.11% Ischaemic  *M =* 12.26 (3.56)  *M =* 6.82 (6.10)  32.24% Impaired | *M =* 70.35 (20.54)  100.00% Male  100.00% Ischaemic  *M =* 10.00 (1.41)  NA  0% Impaired | *t* = 0.24  ꭓ^2^ *=* 0.37  ꭓ^2^ *=* 0.00  *t =* 2.23  NA  ꭓ^2^ *=* 0.71 |
|  | **Complete 6-Month Attention (*N* = 413)** | **Missing 6-Month Attention (*N* = 17)** |  |
| **Demographics**  Age  Sex  Stroke Type  Years of Education  Acute NIHSS Score  Acute Attention Impairment | *M =* 73.79 (12.36)  53.13% Male  83.54% Ischaemic  *M =* 12.29 (3.58)  *M =* 6.76 (6.06)  33.66% Impaired | *M =* 75.34 (16.05)  52.94% Male  100.00% Ischaemic  *M =* 11.24 (2.66)  *M =* 9.22 (7.08)  0% Impaired | *t* = -0.39  ꭓ^2^ *=* 0.00  ꭓ^2^ *=* 2.20  *t =* 1.58  *t =* -1.03  ꭓ^2^ *=* 3.28 |
|  | **Complete 6-Month Numeracy (*N* = 420)** | **Missing 6-Month Numeracy (*N* = 10)** |  |
| **Demographics**  Age  Sex  Stroke Type  Years of Education  Acute NIHSS Score  Acute Numeracy Impairment | *M =* 73.67 (12.54)  54.29% Male  84.05% Ischaemic  *M =* 12.30 (3.53)  *M =* 6.75 (5.94)  20.24% Impaired | *M =* 81.83 (8.01)  20.00% Male  90.00% Ischaemic  *M =* 10.10 (4.09)  *M =* 10.71 (10.81)  0% Impaired | ***t* = -3.13***  ꭓ^2^ *=* 3.34  ꭓ^2^ *=* 0.01  *t =* 1.68  *t =* -0.97  **ꭓ^2^ *=* 7.94**** |
|  | **Complete 6-Month Executive Function (*N* = 400)** | **Missing 6-Month Executive Function (*N* = 30)** |  |
| **Demographics**  Age  Sex  Stroke Type  Years of Education  Acute NIHSS Score  Acute Executive Function Impairment | *M =* 73.59 (12.41)  54.00% Male  84.00% Ischaemic  *M =* 12.32 (3.46)  *M =* 6.78 (6.13)  24.50% Impaired | *M =* 77.39 (13.51)  46.67% Male  86.67% Ischaemic  *M =* 11.24 (4.63)  *M =* 7.76 (5.66)  0% Impaired | *t* = -1.49  ꭓ^2^ *=* 0.34  ꭓ^2^ *=* 0.02  *t =* 1.23  *t =* -0.69  ꭓ^2^ *=* 0.00 |
|  | **Complete 6-Month Praxis (*N* = 403)** | **Missing 6-Month Praxis (*N* = 27)** |  |
| **Demographics**  Age  Sex  Stroke Type  Years of Education  Acute NIHSS Score  Acute Praxis Impairment | *M =* 73.61 (12.45)  53.85% Male  84.12% Ischaemic  *M =* 12.28 (3.59)  *M =* 6.74 (6.08)  19.60% Impaired | *M =* 77.57 (13.01)  48.15% Male  85.19% Ischaemic  *M =* 11.81 (2.99)  *M =* 8.26 (6.30)  0% Impaired | *t* = -1.54  ꭓ^2^ *=* 0.14  ꭓ^2^ *=* 0.00  *t =* 0.76  *t =* -1.02  ꭓ^2^ *=* 0.72 |

**Table S1.** Mean and frequency statistics of demographic, cognitive, and stroke outcome data for participants that had missing information on any of the 6-month domain outcomes compared to those without missing data. There was no missingness at 6-months on the memory domain and hence sensitivity analyses were not conducted. Where NA is listed, this indicates that the participants with missing data did not have corresponding values available to compare to those with complete cases (e.g., the 2 participants with missing data in the language domain did not have acute NIHSS scores).

OCS = Oxford Cognitive Screen; NIHSS = National Institute of Health Stroke Severity; mRS = Modified Rankin Scale

* *p* < 0.05, ** *p* < 0.01, *** *p* < 0.001

In terms acute NIHSS scores, 28.8% (*N* missing = 124 of 430) had missing data. In general, there was no influence of demographic or PSCI status on NIHSS scoress (see *Table S2*)

|  | **Complete Acute NIHSS (*N* = 306)** | **Missing Acute NIHSS (*N* = 124)** |  |
| --- | --- | --- | --- |
| **Demographics**  Age  Sex  Stroke Type  Years of Education  Acute Proportion OCS Tasks Impaired  Acute Language Impairment  Acute Memory Impairment  Acute Attention Impairment  Acute Numeracy Impairment  Acute Executive Impairment  Acute Praxis Impairment | *M =* 74.58 (12.39)  51.63% Male  82.68% Ischaemic  *M =* 12.22 (3.39)  *M =* 0.26 (0.24)  46.08% Impaired  39.22% Impaired  54.58% Impaired  39.22% Impaired  25.16% Impaired  26.47% Impaired | *M =* 72.07 (12.68)  58.06% Male  87.90% Ischaemic  *M =*12.31 (3.94)  *M =* 0.29 (0.26)  43.55% Impaired  40.32% Impaired  45.16% Impaired  46.77% Impaired  29.03% Impaired  25.00% Impaired | *t* = -1.87  ꭓ^2^ *=* 1.22  ꭓ^2^ *=* 1.44  *t =* -0.22  *t* = -0.94  ꭓ^2^ *=* 0.09  ꭓ^2^ *=* 0.02  ꭓ^2^ *=* 1.67  ꭓ^2^ *=* 0.84  ꭓ^2^ *=* 0.04  ꭓ^2^ *=* 1.89 |

**Table S2.** Mean and frequency statistics of demographic, cognitive, and stroke outcome data for participants that had missing information on acute NIHSS scores.

OCS = Oxford Cognitive Screen; NIHSS = National Institute of Health Stroke Severity;

* *p* < 0.05, ** *p* < 0.01, *** *p* < 0.001

***Exploratory Domain-Specific Prediction Models***

At 6-months post-stroke, there was variation in complete outcome data across domains (Language *N*=428; Memory *N*=430; Attention *N=*416; Praxis *N*=403; Numeracy *N*=420; Executive Function *N*=400).

Pooled shrunken coefficients of domain-specific prediction models are shown below in Tables S3 – S8.

|  | **6-Month Language Impairment (binary)** | |
| --- | --- | --- |
|  | **OR [95% CI]** | ***p*-value** |
| **Clinically Relevant Predictors** | | |
| **Intercept** | 1.06 [0.79 – 1.44] | 0.68 |
| **Age at Stroke** | 1.00 [0.99 – 1.01] | 0.07 |
| **Sex** (Female) | 0.97 [0.90 – 1.05] | 0.41 |
| **Years of Education** | 0.99 [0.98 – 1.00] | 0.09 |
| **NIHSS** | 1.00 [0.99 – 1.01] | 0.22 |
| **Stroke Hemisphere**  Right  Bilateral  Undetermined from Scan | 0.91 [0.84 – 0.99]  0.89 [0.77 – 1.03]  0.95 [0.85 – 1.06] | <0.05  0.12  0.36 |
| **First/Recurrent Stroke**  Recurrent Stroke | 0.99 [0.92 – 1.08] | 0.92 |
| **Stroke Type**  Haemorrhagic | 0.98 [0.88 – 1.08] | 0.66 |
| **Acute OCS Language Impairment** | 1.38 [1.28 – 1.49] | <0.001 |
| **Data Driven Predictors** | | |
| **Independence Before Admission**  Carer Support  Family Support | 1.23 [1.07 – 1.41]  1.05 [0.88 – 1.27] | <0.01  0.58 |

**Table S3**. Pooled odds ratios estimates of the language model. Shrinkage was applied to regression odds ratio (OR) estimates using the optimism-adjusted C-Slope obtained via bootstrapping. For the categorical variables stroke hemisphere and independence before admission, the reference categories were left hemisphere stroke and independent prior to admission, respectively.

OCS = Oxford Cognitive Screen; OR = Odds Ratio

|  | **6-Month Memory Impairment (binary)** | |
| --- | --- | --- |
|  | **OR [95% CI]** | ***p*-value** |
| **Clinically Relevant Predictors** | | |
| **Intercept** | 1.13 [0.84 – 1.52] | 0.41 |
| **Age at Stroke** | 1.00 [0.99 – 1.00] | 0.32 |
| **Sex** (Female) | 1.03 [0.95 – 1.11] | 0.48 |
| **Years of Education** | 0.99 [0.98 – 1.00] | 0.09 |
| **NIHSS** | 1.00 [0.99 – 1.01] | 0.68 |
| **Stroke Hemisphere**  Right  Bilateral  Undetermined from Scan | 1.01 [0.93 – 1.10]  0.89 [0.77 – 1.02]  0.99 [0.89 – 1.11] | 0.76  0.10  0.89 |
| **First/Recurrent Stroke**  Recurrent Stroke | 1.02 [0.95 – 1.11] | 0.54 |
| **Stroke Type**  Haemorrhagic | 0.98 [0.89 – 1.08] | 0.68 |
| **Acute OCS Memory Impairment** | 1.29 [1.20 – 1.40] | <0.001 |
| **Data Driven Predictors** | | |
| **Independence Before Admission**  Carer Support  Family Support | 1.34 [1.17 – 1.54]  1.01 [0.85 – 1.21] | <0.001  0.89 |

**Table S4.** Pooled estimates of adjusted beta coefficients and odds ratios of the memory model. Shrinkage was applied to regression odds ratio (OR) estimates using the optimism-adjusted C-Slope obtained via bootstrapping. For the categorical variables stroke hemisphere and independence before admission, the reference categories were left hemisphere stroke and independent prior to admission, respectively.

OCS = Oxford Cognitive Screen; OR = Odds Ratio

|  | **6-Month Attention Impairment (binary)** | |
| --- | --- | --- |
|  | **OR [95% CI]** | ***p*-value** |
| **Clinically Relevant Predictors** | | |
| **Intercept** | 0.75 [0.56 – 1.01] | 0.07 |
| **Age at Stroke** | 1.01 [1.00 – 1.01] | <0.001 |
| **Sex** (Female) | 1.04 [0.96 – 1.12] | 0.37 |
| **Years of Education** | 0.99 [0.98 – 0.99] | <0.05 |
| **NIHSS** | 1.00 [0.99 – 1.01] | 0.48 |
| **Stroke Hemisphere**  Right  Bilateral  Undetermined from Scan | 1.13 [1.04 – 1.23]  1.01 [0.87 – 1.17]  0.96 [0.86 – 1.07] | <0.01  0.93  0.45 |
| **First/Recurrent Stroke**  Recurrent Stroke | 1.08 [0.99 – 1.18] | 0.10 |
| **Stroke Type**  Haemorrhagic | 0.96 [0.87 – 1.07] | 0.45 |
| **Acute OCS Attention Impairment** | 1.20 [1.11 – 1.30] | <0.001 |
| **Data Driven Predictors** | | |
| **Charlson Comorbidity Index** | 1.02 [0.99 – 1.06] | 0.18 |

**Table S5.** Pooled estimates of adjusted beta coefficients and odds ratios of the attention model. Shrinkage was applied to regression odds ratio (OR) estimates using the optimism-adjusted C-Slope obtained via bootstrapping. For the categorical variables stroke hemisphere and independence before admission, the reference categories were left hemisphere stroke and independent prior to admission, respectively.

OCS = Oxford Cognitive Screen; OR = Odds Ratio

|  | **6-Month Numeracy Impairment (binary)** | |
| --- | --- | --- |
|  | **OR [95% CI]** | ***p*-value** |
| **Clinically Relevant Predictors** | | |
| **Intercept** | 1.20 [0.92 – 1.56] | 0.18 |
| **Age at Stroke** | 0.99 [0.99 – 1.00] | 0.37 |
| **Sex** (Female) | 1.06 [0.99 – 1.13] | 0.11 |
| **Years of Education** | 0.99 [0.98 – 1.01] | 0.51 |
| **NIHSS** | 1.00 [0.99 – 1.01] | 0.23 |
| **Stroke Hemisphere**  Right  Bilateral  Undetermined from Scan | 0.99 [0.92 – 1.06]  0.93 [0.82 – 1.06]  1.07 [0.97 – 1.17] | 0.75  0.26  0.20 |
| **First/Recurrent Stroke**  Recurrent Stroke | 1.03 [0.96 – 1.10] | 0.43 |
| **Stroke Type**  Haemorrhagic | 0.93 [0.85 – 1.01] | 0.10 |
| **Acute OCS Numeracy Impairment** | 1.20 [1.12 – 1.29] | <0.001 |

**Table S6.** Pooled estimates of adjusted beta coefficients and odds ratios of the numeracy model. Shrinkage was applied to regression odds ratio (OR) estimates using the optimism-adjusted C-Slope obtained via bootstrapping. For the categorical variables stroke hemisphere and independence before admission, the reference categories were left hemisphere stroke and independent prior to admission, respectively.

OCS = Oxford Cognitive Screen; OR = Odds Ratio

|  | **6-Month Executive Function Impairment (binary)** | |
| --- | --- | --- |
|  | **OR [95% CI]** | ***p*-value** |
| **Clinically Relevant Predictors** | | |
| **Intercept** | 0.80 [0.61 – 1.05] | 0.11 |
| **Age at Stroke** | 1.00 [1.00 – 1.01] | <0.01 |
| **Sex** (Female) | 1.09 [1.02 – 1.17] | <0.05 |
| **Years of Education** | 0.99 [0.98 – 1.01] | 0.37 |
| **NIHSS** | 1.00 [0.99 – 1.01] | 0.13 |
| **Stroke Hemisphere**  Right  Bilateral  Undetermined from Scan | 0.99 [0.92 – 1.08]  1.00 [0.88 – 1.15]  0.96 [0.87 – 1.07] | 0.98  0.95  0.46 |
| **First/Recurrent Stroke**  Recurrent Stroke | 1.02 [0.94 – 1.10] | 0.68 |
| **Stroke Type**  Haemorrhagic | 0.91 [0.83 – 1.00] | 0.05 |
| **Acute OCS Executive Function Impairment** | 1.16 [1.07 – 1.25] | <0.001 |
| **Data Driven Predictors** | | |
| **Charlson Comorbidity Index** | 1.02 [0.99 – 1.06] | 0.15 |

**Table S7.** Pooled estimates of adjusted beta coefficients and odds ratios of the executive function model. Shrinkage was applied to regression odds ratio (OR) estimates using the optimism-adjusted C-Slope obtained via bootstrapping. For the categorical variables stroke hemisphere and independence before admission, the reference categories were left hemisphere stroke and independent prior to admission, respectively.

OCS = Oxford Cognitive Screen; OR = Odds Ratio

|  | **6-Month Praxis Impairment (binary)** | |
| --- | --- | --- |
|  | **OR [95% CI]** | ***p*-value** |
| **Clinically Relevant Predictors** | | |
| **Intercept** | 0.94 [0.76 – 1.16] | 0.56 |
| **Age at Stroke** | 1.00 [1.00 – 1.01] | <0.01 |
| **Sex** (Female) | 0.95 [0.89 – 0.99] | 0.05 |
| **Years of Education** | 0.99 [0.99 – 1.01] | 0.66 |
| **NIHSS** | 1.00 [0.99 – 1.01] | 0.70 |
| **Stroke Hemisphere**  Right  Bilateral  Undetermined from Scan | 0.97 [0.91 – 1.03]  0.88 [0.80 – 0.98]  0.99 [0.92 – 1.08] | 0.27  0.02  0.97 |
| **First/Recurrent Stroke**  Recurrent Stroke | 1.02 [0.96 – 1.08] | 0.46 |
| **Stroke Type**  Haemorrhagic | 1.02 [0.95 – 1.10] | 0.53 |
| **Acute OCS Praxis Impairment** | 1.07 [1.01 – 1.14] | <0.05 |
| **Data Driven Predictors** | | |
| **Independence Before Admission**  Carer Support  Family Support | 0.92 [0.83 – 1.02]  1.11 [0.97 – 1.27] | 0.09  0.14 |

**Table S8.** Pooled estimates of adjusted beta coefficients and odds ratios of the praxis model. Shrinkage was applied to regression odds ratio (OR) estimates using the optimism-adjusted C-Slope obtained via bootstrapping. For the categorical variables stroke hemisphere and independence before admission, the reference categories were left hemisphere stroke and independent prior to admission, respectively.

OCS = Oxford Cognitive Screen; OR = Odds Ratio

**Domain-Specific Prediction Model Results**

The below summarises the key model performance statistics across each cognitive domain. Methods are detailed in the main manuscript.

*Language Model*

In the final multivariable language model, higher age (pooled OR=1.00 [95% CI=0.99–1.01]), years of education (pooled OR=0.99 [95% CI=0.98–1.00]), left hemisphere stroke (pooled OR=0.91 [95% CI=0.84–0.99]) and acute language impairments (pooled OR = 1.38 [95% CI=1.28–1.49]) were most strongly associated with 6-month language impairment. Of the data-driven predictors, only requiring carer support prior to admission was retained (pooled OR=1.23 [95% CI=1.07–1.41]).

The final optimism-adjusted language model showed good performance (C-Statistic=0.77 [95% CI=0.72–0.81]; C-Slope=0.91 [95% CI=0.75–1.07]; CITL= -0.98 [95% CI= -1.18– -0.78]; Brier Score=0.10 [95% CI=0.08–0.11]; Nagelkerke’s R^2^=0.25), with no notable predictor selection differences in complete case data.

*Memory Model*

In the final multivariable memory model, acute memory impairments (pooled OR=1.29 [95% CI=1.20–1.40]) predicted likelihood of 6-month memory impairment *(p*<0.10). Of the data-driven predictors, only requiring carer support prior to admission was retained OR=1.36 [95% CI=1.17–1.54]).

The final optimism-adjusted memory model showed acceptable to good performance (C-Statistic=0.70 [95% CI=0.65–0.75]; C-Slope=0.88 [95% CI=0.71–1.06]; CITL= -0.92 [95% CI= -1.13– -0.72]; Brier Score=0.10 [95% CI=0.08–0.11]; Nagelkerke’s R^2^=0.16), with no differences in predictor selection in complete case data.

*Attention Model*

For the final multivariable attention model, higher age (pooled OR=1.01 [95% CI=1.00–1.01]), lower number of years of education (pooled OR=0.99 [95% CI=0.98–0.99]), right hemisphere lesions (pooled OR=1.13 [95% CI=1.04–1.23]), recurrent stroke (pooled OR=1.08 [95% CI=0.99–1.18]) and acute attention impairments (pooled OR=1.20 [95% CI=1.11–1.30]) had the strongest associations with likelihood of 6-month attention impairment, with only greater CCI scores retained from the data-driven predictors (pooled OR=1.02 [95% CI=0.99–1.06]).

The final optimism-adjusted attention model showed acceptable to good performance (C-Statistic=0.73 [95% CI=0.68–0.77]; C-Slope=0.88 [95% CI=0.70–1.06]; CITL= -1.27 [95% CI= -1.47– -1.06]; Brier Score=0.07 [95% CI=0.05–0.08]; Nagelkerke’s R^2^=0.19), with no predictor selection differences in complete cases.

*Numeracy Model*

In the final multivariable numeracy model, non-haemorrhagic stroke (pooled OR=0.93 [95% CI=0.85–1.01]) and acute numeracy impairments (pooled OR=1.20 [95% CI=1.12–1.29]) demonstrated the strongest predictive value toward likelihood of 6-month numeracy impairments. No data-driven predictors were retained (*p*s>0.10).

The final optimism-adjusted numeracy model showed acceptable performance (C-Statistic=0.69 [95% CI=0.63–0.75]; C-Slope=0.86 [95% CI=0.59–1.10]; CITL= -1.37 [95% CI= -1.61– -1.13]; Brier Score=0.08 [95% CI=0.07–0.10]; Nagelkerke’s R^2^=0.11), with no differences in predictor selection in complete cases.

*Executive Function Model*

In the final multivariable executive function model, female sex (pooled OR=1.09 [95% CI=1.02–1.17]), non-haemorrhagic stroke (pooled OR=0.91 [95% CI=0.83–1.00]), and acute executive function impairments (pooled OR=1.16 [95% CI=1.07–1.25]) demonstrated the strongest associations with 6-month executive function impairments. Of the data-driven predictors, only greater CCI scores were retained (pooled OR=1.02 [95% CI=0.99–1.06]).

The final optimism-adjusted executive function model showed acceptable performance (C-Statistic=0.71 [95% CI=0.65–0.76]; C-Slope=0.82 [95% CI=0.58–1.06]; CITL= -1.55 [95% CI= -1.78– -1.32]; Brier Score=0.10 [95% CI=0.07–0.11]; Nagelkerke’s R^2^=0.13), with no differences in predictor selection in complete case data.

*Praxis Model*

In the final multivariable praxis model, greater age at time of stroke (pooled OR=1.00 [95% CI=1.00–1.01]), male sex (pooled OR=0.95 [95% CI=0.89–0.99]), bilateral hemisphere lesions (pooled OR=0.88 [95% CI=0.80–0.98]), and acute praxis impairments (pooled OR=1.07 [95% CI=1.01–1.14]) were most strongly associated with likelihood of 6-month praxis impairments. Of the data-driven predictors, only requiring carer support prior to admission was retained (pooled OR=0.92 [95% CI=0.83–1.02]).

The final optimism-adjusted praxis model showed poor to acceptable performance (C-Statistic=0.60 [95% CI=0.54–0.66]; C-Slope=0.69 [95% CI=0.37–1.01]; CITL= -1.61 [95% CI= -1.86– -1.37]; Brier Score=0.07 [95% CI=0.04–0.08]; Nagelkerke’s R^2^=0.01). No predictor selection differences were observed in complete cases.

All domain-specific model performance measures are in Table S9 below. Unadjusted relationships between predictor and outcome variables are in *Tables S11–S16.* Domain-specific calibration plots are in Figures S1 (complete cases) and S2 (pooled imputed data)*.*

| **Model** | **Estimate** | **Model**  **Performance**  **[95% CI]** | **Average**  **Optimism**  **[95% CI]** | **Optimism-Adjusted Performance**  **[95% CI]** |
| --- | --- | --- | --- | --- |
| **Language** | C-Statistic  C-Slope  CITL  Brier Score  E/O  Nagelkerke’s R^2^ | 0.80 [0.75–0.84]  1.01 [0.84–1.17]  0.0001 [0.0001–0.0001]  0.16 [0.14–0.18]  1 [1–1]  0.32 | 0.03 [0.03–0.04]  0.10 [0.09–0.11]  0.98 [0.78–1.89]  0.06 [0.06–0.07]  0.00 [-0.12–0.12]  0.07 [0.07–0.08] | 0.77 [0.72–0.81]  0.91 [0.75–1.07]  -0.98 [-1.18– -0.78]  0.10 [0.08–0.11]  1.00 [0.88–1.12]  0.25 |
| **Memory** | C-Statistic  C-Slope  CITL  Brier Score  E/O  Nagelkerke’s R^2^ | 0.75 [0.70–0.80]  1.00 [0.81–1.19]  -0.0001 [-0.0001- -0.0001]  0.17 [0.15–0.19]  1 [1–1]  0.26 | 0.05 [0.05–0.05]  0.13 [0.12–0.13]  0.92 [1.13–0.72]  0.08 [0.07–0.08]  0.001 [-0.13–0.13]  0.09 [0.08–0.09] | 0.70 [0.65–0.75]  0.88 [0.71–1.06]  -0.92 [-1.13– -0.72]  0.10 [0.08–0.11]  0.99 [0.87–1.13]  0.16 |
| **Attention** | C-Statistic  C-Slope  CITL  Brier Score  E/O  Nagelkerke’s R^2^ | 0.78 [0.73–0.82]  1.01 [0.83–1.19]  -0.0001 [-0.0001– -0.0001]  0.14 [0.12–0.16]  1 [1–1]  0.29 | 0.05 [0.05–0.05]  0.13 [0.12–0.14]  1.27 [1.06–1.47]  0.08 [0.07–0.08]  0.00 [-0.12–0.12]  0.09 [0.09–0.09] | 0.73 [0.68–0.77]  0.88 [0.70–1.06]  -1.27 [-1.47– -1.06]  0.07 [0.05–0.08]  1.00 [0.88–1.12]  0.19 |
| **Numeracy** | C-Statistic  C-Slope  CITL  Brier Score  E/O  Nagelkerke’s R^2^ | 0.74 [0.68–0.79]  1.01 [0.75–1.27]  -0.0001 [-0.0001- -0.0001]  0.14 [0.12–0.16]  1 [1–1]  0.18 | 0.05 [0.04–0.05]  0.16 [0.15–0.16]  1.37 [1.13–1.61]  0.06 [0.06–0.06]  0.00 [-0.18–0.18]  0.07 [0.07–0.07] | 0.69 [0.63–0.75]  0.86 [0.59–1.10]  -1.37 [-1.61– -1.13]  0.08 [0.07–0.10]  1.00 [0.82–1.18]  0.11 |
| **Executive Function** | C-Statistic  C-Slope  CITL  Brier Score  E/O  Nagelkerke’s R^2^ | 0.75 [0.69–0.80]  0.99 [0.74–1.24]  -0.0001 [-0.03–0.03]  0.14 [0.14–0.18]  1 [1–1]  0.19 | 0.04 [0.03–0.04]  0.17 [0.16–0.17]  0.98 [0.97–0.99]  0.06 [0.06–0.07]  0.24 [0.03–0.44]  0.06 [0.06–0.07] | 0.71 [0.65–0.76]  0.82 [0.58–1.06]  -1.55 [-1.78– -1.32]  0.10 [0.07–0.11]  1.24 [1.03–1.44]  0.13 |
| **Praxis** | C-Statistic  C-Slope  CITL  Brier Score  E/O  Nagelkerke’s R^2^ | 0.69 [0.62–0.75]  0.99 [0.66–1.32]  -0.0001 [-0.03–0.03]  0.16 [0.12–0.17]  1 [1–1]  0.13 | 0.09 [0.08–0.09]  0.30 [0.29–0.31]  1.61 [1.83–1.34]  0.09 [0.08–0.09]  0.00 [-0.19–0.19]  0.11 [0.11–0.11] | 0.60 [0.54–0.66]  0.69 [0.37–1.01]  -1.61 [-1.86– -1.37]  0.07 [0.04–0.08]  1.00 [0.81–1.20]  0.01 |

**Table S9.** Performance metrics all final models pooled across 20 imputed datasets.

CITL=Calibration-in-the-large; E/O=Expected:Observed Ratio; C-Slope=Calibration Slope

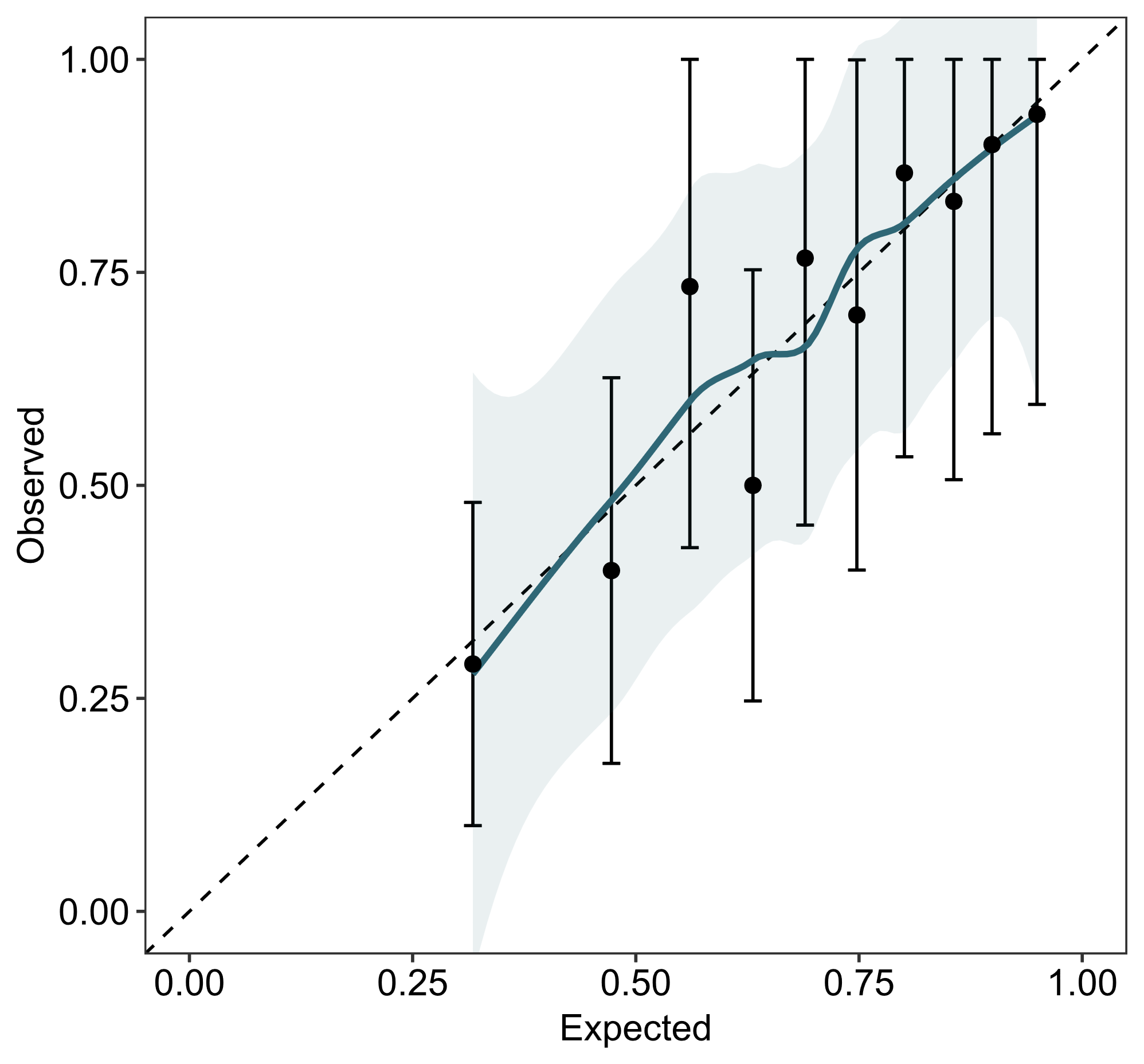

**Figure S1**. Calibration plot of any 6-month PSCI (0=no impairment, 1=any impairment) in complete case data*.*

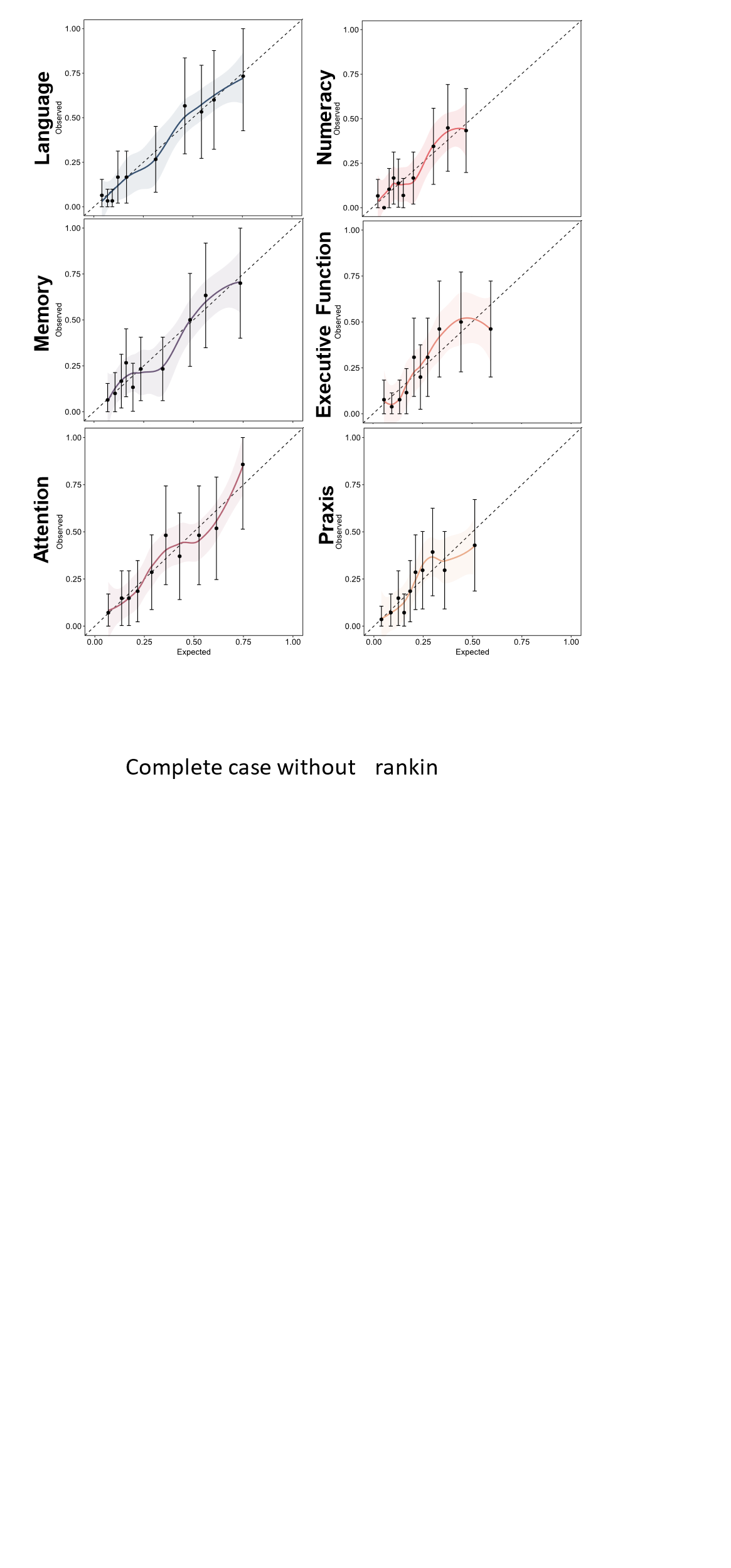

**Figure S2**. Calibration plots of 6-month language, memory, attention, numeracy, executive function, and praxis impairments using complete cases. Impairments were modelled dichotomously (0 = no impairment, 1 = any impairment) risk prediction model with a Loess smoother in pooled imputed data across complete case data. Points indicate risk groups by decile with 95% confidence intervals. Predicted probabilities are on the x-axis, while observed probabilities are on the y-axis. Values closer to the reference line (dashed) indicate better model fit.

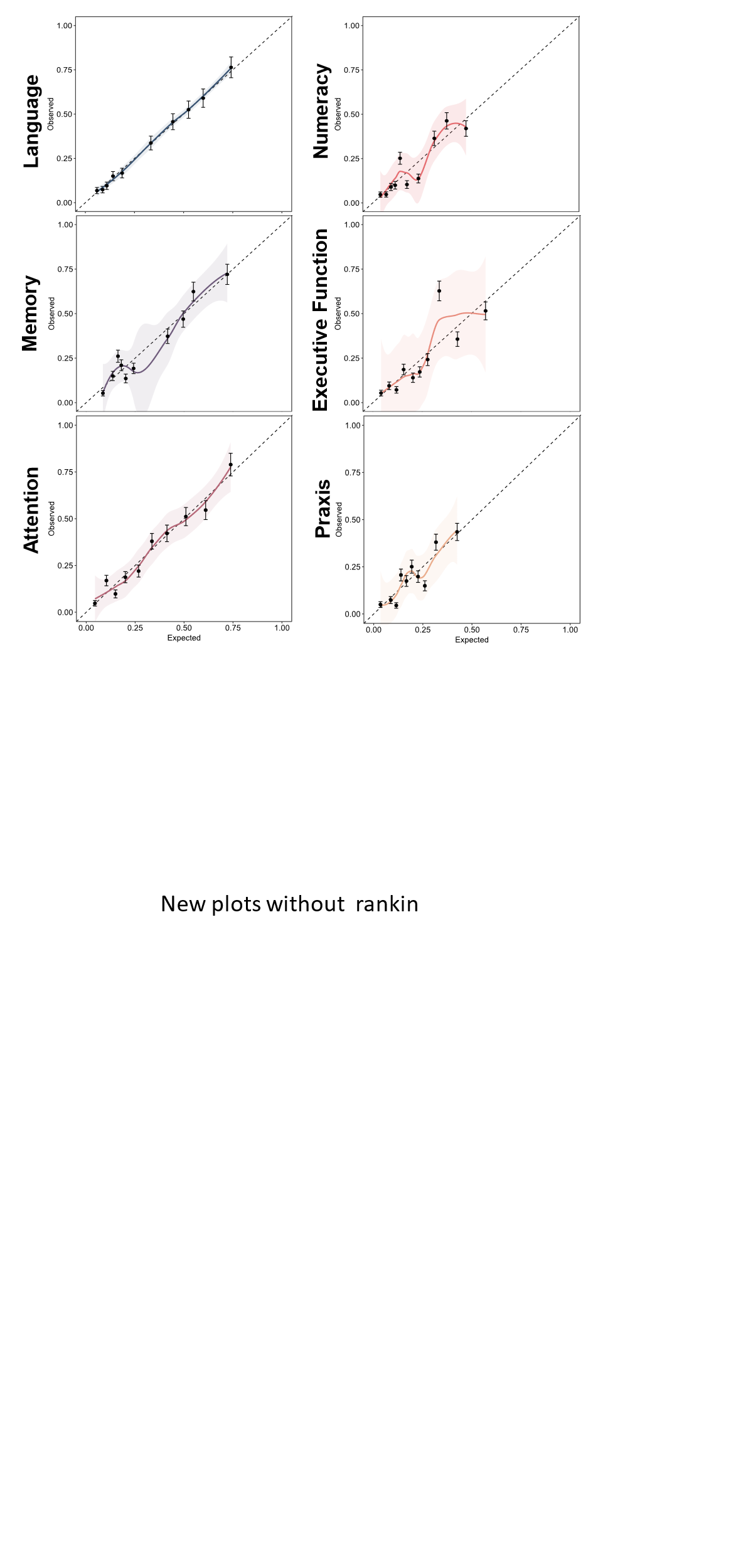

**Figure S2**. Calibration plots of 6-month domain-specific impairments (0 = no impairment, 1 = any impairment) across 20 imputed datasets.

***External validation of Domain-Specific Models***

Below details external validation performance of domain-specific models using the OCS-Care dataset. Methods for obtaining performance metrics are in the main manuscript.

| **Model** | **Estimate** | **Model**  **Performance**  **[95% CI]** |
| --- | --- | --- |
| **Language** | C-Statistic  C-Slope  CITL  Nagelkerke’s R^2^ | 0.77 [0.69–0.84]  6.10 [4.08–8.11]  -2.84 [-3.49– -2.19]  0.23 |
| **Memory** | C-Statistic  C-Slope  CITL  Nagelkerke’s R^2^ | 0.69 [0.60–0.77]  5.29 [2.84–7.74]  -2.60 [-3.29–1.92]  0.11 |
| **Attention** | C-Statistic  C-Slope  CITL  Nagelkerke’s R^2^ | 0.75 [0.66–0.82]  5.95 [3.59–8.31]  -2.76 [-3.47– -2.05]  0.20 |
| **Numeracy** | C-Statistic  C-Slope  CITL  Nagelkerke’s R^2^ | 0.79 [0.70–0.86]  12.06 [8.05–16.08]  -3.51 [-4.36– -2.66]  0.27 |
| **Executive Function** | C-Statistic  C-Slope  CITL  Nagelkerke’s R^2^ | 0.74 [0.62–0.83]  8.54 [4.42– 12.65]  -3.39 [-4.27– -2.51]  0.15 |
| **Praxis** | C-Statistic  C-Slope  CITL  Nagelkerke’s R^2^ | 0.66 [0.55–0.75]  11.67 [3.60–19.73]  -3.44 [-4.59– -2.29]  0.08 |

**Table S10.** Model performance following external validation across 20 imputed datasets using OCS-Care data.

CITL=Calibration-in-the-large; C-Slope=Calibration Slope

| **Overall PSCI Model Predictors** | **Summary Statistics** | | **Adjusted R^2^** | **Pooled Adjusted R^2^** |
| --- | --- | --- | --- | --- |
|  | **No 6 Months OCS Impairment** | **6 Month OCS Impairment** |  |  |
| **Clinically Relevant Predictors** | | | | |
| **Age at Stroke** – M (SD) | 69.77 (13.06) | 75.73 (11.81) | **0.05 (*p* < 0.001)** | **0.05 (*p* < 0.001)** |
| **Sex** (n, %)  Male  Female | 78 (58%)  57 (42%) | 152 (52%)  143 (48%) | 0.004 (*p =* 0.10*)* | 0.004 (*p =* 0.10*)* |
| **Years of Education** – M (SD) | 13.13 (3.99) | 11.84 (3.26) | **0.04 (*p* < 0.001)** | **0.04 (*p* < 0.001)** |
| **NIHSS** (median) | 4 | 6 | **0.02 (*p <* 0.01)** | **0.02 (*p <* 0.01)** |
| **Stroke Hemisphere** (n, %)  Left  Right  Bilateral  Undetermined from Scan | 40 (30%)  51 (38%)  18 (13%)  26 (19%) | 113 (38%)  117 (40%)  16 (5%)  49 (17%) | **0.01 (*p <* 0.05)** | **0.01 (*p <* 0.05)** |
| **First/Recurrent Stroke** (n, %)  First Stroke  Recurrent Stroke | 103 (76%)  32 (24%) | 189 (64%)  106 (36%) | 0.01 (*p =* 0.10) | 0.01 (*p =* 0.08) |
| **Stroke Type** (n, %)  Ischaemic  Haemorrhagic  Mixed | 110 (81%)  25 (19%)  0 (0%) | 252 (85%)  40 (14%)  3 (1%) | 0.01 (*p* = 0.10) | 0.01 (*p* = 0.06) |
| **Data Driven Predictors** | | | | |
| **Length of Stay** – median | 6 | 10 | **0.02 (*p* < 0.01)** | **0.02 (*p* < 0.01)** |
| **Acute Mood Difficulties** (n, %)  Yes  No | 104 (77.0%)  31 (23.0%) | 221 (75.0%)  72 (25.0%) | -0.002 (*p* = 0.75) | 0.001 (*p =* 0.74) |
| **Independence Before Admission** (n, %)  Independent  Carer Support  Family Support | 128 (95%)  4 (3%)  3 (2%) | 248 (84%)  32 (11%)  15 (5%) | **0.06 (*p* < 0.001)** | **0.06 (*p* < 0.001)** |
| **Charlson Comorbidity Index** – M (SD) |  |  | 0.01 (*p* = 0.06) | 0.01 (*p* = 0.06) |
| **Acute Proportion OCS Tasks Impaired** – M (SD) | 0.13 (0.20) | 0.33 (0.24) | **0.30 (*p* < 0.001)** | **0.30 (*p* < 0.001)** |

**Table S11.** Descriptive statistics of model predictors between those with (*N* = 295) and without (*N* = 135) a 6-month PSCI impairment.

| **Language Model Predictors** | **Summary Statistics** | | **Odds Ratio** | **Pooled Odds Ratio** |
| --- | --- | --- | --- | --- |
|  | **No 6 Month Language Impairment** | **6 Month Language Impairment** |  |  |
| **Clinically Relevant Predictors** | | | | |
| **Age at Stroke** – M (SD) | 72.71 (12.12) | 76.30 (12.96) | **1.04 (*p* < 0.01)** | **1.03** |
| **Sex** (n, %)  Male  Female | 154 (53.0%)  136 (47.0%) | 74 (54.0%)  64 (46.0%) | 1.21 (*p =* 0.44*)* | 0.97 |
| **Years of Education** – M (SD) | 12.68 (3.64) | 11.39 (3.21) | **0.85 (*p* < 0.001)** | **0.89** |
| **NIHSS** (median) | 4 | 6 | 1.04 (*p =* 0.06) | 1.05 |
| **Stroke Hemisphere** (n, %)  Left  Right  Bilateral  Undetermined from Scan | 87 (30.0%)  126 (43.0%)  27 (27.0%)  50 (17.0%) | 66 (48.0%)  42 (30.0%)  5 (4.0%)  25 (18.0%) | **0.49 (*p <* 0.05)**  **0.25 (*p <* 0.05)**  0.82 (*p =* 0.58) | **0.44**  **0.24**  0.66 |
| **First/Recurrent Stroke** (n, %)  First Stroke  Recurrent Stroke | 199 (69.0%)  91 (31.0%) | 92 (67.0%)  46 (33.0%) | 0.90 (*p =* 0.71) | 1.09 |
| **Stroke Type** (n, %)  Ischaemic  Haemorrhagic | 240 (83.0%)  50 (17.0%) | 120 (87.0%)  18 (13.0%) | 0.51 (*p =* 0.06) | 0.72 |
| **Length of Stay** – median | 8 | 10 | 1.02 (*p* = 0.14) | 1.02 |
| **Acute Mood Difficulties** (n, %)  Yes  No | 221 (76.0%)  68 (24.0%) | 103 (75.0%)  34 (25.0%) | 1.45 (*p* = 0.18) | 1.07 (*p =* 0.78) |
| **Independence Before Admission** (n, %)  Independent  Carer Support  Family Support | 266 (92.0%)  15 (5.0%)  9 (3.0%) | 108 (78.0%)  21 (15.0%)  9 (7.0%) | **9.70 (*p* < 0.001)**  1.72 (*p =* 0.32) | **3.45**  2.46 |
| **Charlson Comorbidity Index** – M (SD) | 1.33 (1.23) | 1.46 (1.27) | 1.09 (*p* = 0.37) | 1.09 |
| **Acute Language Impairment** (n, %)  No Impairment  Impairment | 201 (70.0%)  88 (30.0%) | 33 (24.0%)  105 (76.0%) | **6.63 (*p* < 0.001)** | **7.30** |

**Table S12.** Descriptive statistics of model predictors between those with (*N* = 138) and without (*N* = 290) a 6-month language impairment.

| **Memory Model Predictors** | **Summary Statistics** | | **Odds Ratio** | **Pooled Odds Ratio** |
| --- | --- | --- | --- | --- |
|  | **No 6 Month Memory Impairment** | **6 Month Memory Impairment** |  |  |
| **Clinically Relevant Predictors** | | | | |
| **Age at Stroke** – M (SD) | 72.60 (12.80) | 76.54 (11.45) | **1.03 (*p* < 0.01)** | **1.03** |
| **Sex** (n, %)  Male  Female | 163 (56.0%)  130 (44.0%) | 67 (49.0%)  70 (51.0%) | 1.21 (*p =* 0.44*)* | 1.31 |
| **Years of Education** – M (SD) | 12.64 (3.66) | 11.39 (3.16) | **0.82 (*p* < 0.001)** | **0.89** |
| **NIHSS** (median) | 5 | 6 | 1.03 (*p =* 0.15) | 1.03 |
| **Stroke Hemisphere** (n, %)  Left  Right  Bilateral  Undetermined from Scan | 99 (34.0%)  116 (40.0%)  28 (10.0%)  50 (17.0%) | 54 (39.0%)  52 (38.0%)  6 (4.0%)  25 (18.0%) | 0.80 (*p =* 0.44)  0.46 (*p =* 0.19)  0.81 (*p =* 0.57) | 0.82  0.39  0.92 |
| **First/Recurrent Stroke** (n, %)  First Stroke  Recurrent Stroke | 205 (70.0%)  88 (30.0%) | 87 (64.0%)  50 (36.0%) | 0.99 (*p =* 0.97) | 1.34 |
| **Stroke Type** (n, %)  Ischaemic  Haemorrhagic | 243 (83.0%)  50 (17.0%) | 119 (87.0%)  18 (13.0%) | 0.55 (*p =* 0.09) | 0.74 |
| **Length of Stay** – median | 8 | 10 | **1.04 (*p <* 0.01)** | **1.03** |
| **Independence Before Admission** (n, %)  Independent  Carer Support  Family Support | 271 (92.0%)  10 (3.0%)  12 (4.0%) | 105 (77.0%)  26 (19.0%)  6 (4.0%) | **7.66 (*p* < 0.001)**  1.82 (*p =* 0.27) | **6.71**  1.29 |
| **Acute Mood Difficulties** (n, %)  Yes  No | 221 (76.0%)  68 (24.0%) | 103 (75.0%)  34 (25.0%) | 1.45 (*p* = 0.18) | 1.07 |
| **Charlson Comorbidity Index** – M (SD) | 1.35 (1.24) | 1.47 (1.35) | 1.08 (*p* = 0.43) | 1.08 |
| **Acute Memory Impairment** (n, %)  No Impairment  Impairment | 210 (72.0%)  81 (28.0%) | 48 (35.0%)  89 (65.0%) | **5.19 (*p* < 0.001)** | **4.76** |

**Table S13.** Descriptive statistics of model predictors between those with (*N* = 137) and without (*N* = 293) a 6-month memory impairment.

| **Attention Model Predictors** | **Summary Statistics** | | **Odds Ratio** | **Pooled Odds Ratio** |
| --- | --- | --- | --- | --- |
|  | **No Attention Impairment at 6 Months** | **Attention Impairment at 6 months** |  |  |
| **Clinically Relevant Predictors** | | | | |
| **Age at Stroke** – M (SD) | 71.41 (12.79) | 78.50 (9.95) | **1.05 (*p* < 0.01)** | **1.06** |
| **Sex** (n, %)  Male  Female | 157 (57.0%)  117 (43.0%) | 64 (46.0%)  75 (54.0%) | **2.02 (*p* < 0.05)** | **1.57** |
| **Years of Education** – M (SD) | 12.70 (3.76) | 11.49 (3.06) | **0.90 (*p* < 0.05)** | **0.90** |
| **NIHSS** (median) | 4 | 6 | 1.03 (*p =* 0.15) | 1.01 |
| **Stroke Hemisphere** (n, %)  Left  Right  Bilateral  Undetermined from Scan | 108 (39.0%)  90 (33.0%)  23 (8.0%)  53 (19.0%) | 37 (27.0%)  75 (54.0%)  9 (6.0%)  18 (13.0%) | **3.64 (*p <* 0.001)**  1.17 (*p =* 0.79)  0.65 (*p =* 0.41) | **2.43**  1.14  0.99 |
| **First/Recurrent Stroke** (n, %)  First Stroke  Recurrent Stroke | 200 (73.0%)  74 (27.0%) | 81 (58.0%)  58 (42.0%) | 1.26 (*p =* 0.45) | 1.94 |
| **Stroke Type** (n, %)  Ischaemic  Haemorrhagic | 226 (82.0%)  48 (18.0%) | 119 (86.0%)  20 (14.0%) | 0.77 (*p =* 0.49) | 0.79 |
| **Data Driven Predictors** | | | | |
| **Length of Stay** – median | 7 | 10 | 1.01 (*p* = 0.66) | 1.01 |
| **Independence Before Admission** (n, %)  Independent  Carer Support  Family Support | 247 (90.0%)  15 (5.0%)  12 (5.0%) | 116 (83.0%)  18 (13.0%)  5 (4.0%) | **4.16 (*p* < 0.05)**  1.39 (*p =* 0.62) | **2.56**  0.89 |
| **Acute Mood Difficulties** (n, %)  Yes  No | 168 (62.0%)  125 (38.0%) | 44 (54.0%)  52 (46.0%) | 1.57 (*p* = 0.16) | 1.03 |
| **Charlson Comorbidity Index** – M (SD) | 1.22 (1.13) | 1.69 (1.42) | **1.24 (*p =* 0.05)** | **1.34** |
| **Acute Attention Impairment** (n, %)  No Impairment  Impairment | 140 (55.0%)  116 (45.0%) | 27 (22.0%)  98 (78.0%) | **4.69 (*p* < 0.001)** | **3.47** |

**Table S14.** Descriptive statistics of model predictors between those with (*N* = 139) and without (*N* = 274) a 6-month attention impairment.

| **Number Model Predictors** | **Summary Statistics** | | **Odds Ratio** | **Pooled Odds Ratio** |
| --- | --- | --- | --- | --- |
|  | **No Numeracy Impairment at 6 Months** | **Numeracy Impairment at 6 months** |  |  |
| **Clinically Relevant Predictors** | | | | |
| **Age at Stroke** – M (SD) | 73.66 (12.33) | 73.68 (13.41) | 0.99 (*p =* 0.76) | 1.00 |
| **Sex** (n, %)  Male  Female | 189 (56.0%)  146 (44.0%) | 39 (46.0%)  46 (54.0%) | 1.53 (*p =* 0.17) | 1.53 |
| **Years of Education** – M (SD) | 12.50 (3.63) | 11.49 (2.97) | **0.88 (*p* < 0.05)** | **0.91** |
| **NIHSS** (median) | 5 | 6 | 1.04 (*p =* 0.11) | **1.04** |
| **Stroke Hemisphere** (n, %)  Left  Right  Bilateral  Undetermined from Scan | 116 (35.0%)  133 (40.0%)  31 (9.0%)  55 (16.0%) | 34 (40.0%)  30 (35.0%)  3 (4.0%)  18 (21.0%) | 0.80 (*p =* 0.51)  0.39 (*p =* 0.24)  1.23 (*p =* 0.61) | 0.77  0.33  1.12 |
| **First/Recurrent Stroke** (n, %)  First Stroke  Recurrent Stroke | 229 (68.0%)  106 (32.0%) | 56 (66.0%)  29 (34.0%) | 0.90 (*p =* 0.74) | 1.12 |
| **Stroke Type** (n, %)  Ischaemic  Haemorrhagic | 275 (82.0%)  60 (18.0%) | 78 (92.0%)  8 (7.0%) | **0.30 (*p <* 0.05)** | **0.41** |
| **Length of Stay** – median | 8 | 10 | 1.00 (*p* = 0.94) | 1.00 |
| **Independence Prior to Admission** (n, %)  Independent  Carer Support  Family Support | 301 (90.0%)  21 (6.0%)  4 (13.0%) | 70 (82.0%)  11 (13.0%)  4 (5.0%) | 1.87 (*p =* 0.26)  1.79 (*p =* 0.34) | **2.25**  1.32 |
| **Acute Mood Difficulties** (n, %)  Yes  No | 255 (77.0%)  78 (23.0%) | 63 (74.0%)  22 (26.0%) | 1.83 (*p* = 0.06) | 1.14 |
| **Charlson Comorbidity Index** – M (SD) | 1.38 (1.27) | 1.46 (1.33) | 1.10 (*p =* 0.41) | 1.05 |
| **Acute Numeracy Impairment** (n, %)  No Impairment  Impairment | 222 (67.0%)  111 (33.0%) | 27 (32.0%)  58 (68.0%) | **4.41 (*p* < 0.001)** | **4.27** |

**Table S15.** Descriptive statistics of model predictors between those with (*N* = 85) and without (*N* = 335) a 6-month numeracy impairment.

| **Executive Function Model Predictors** | **Summary Statistics** | | **Odds Ratio** | **Pooled Odds Ratio** |
| --- | --- | --- | --- | --- |
|  | **No Exec Impairment at 6 Months** | **Exec Impairment at 6 months** |  |  |
| **Clinically Relevant Predictors** | | | | |
| **Age at Stroke** – M (SD) | 72.19 (12.85) | 77.90 (9.82) | **1.04 (*p* <0.01)** | 1.05 |
| **Sex** (n, %)  Male  Female | 176 (58.0%)  126 (42.0%) | 40 (41.0%)  58 (59.0%) | **1.99 (*p* <0.05)** | 2.03 |
| **Years of Education** – M (SD) | 12.54 (3.60) | 11.65 (2.88) | 0.94 (*p* = 0.22) | 0.92 |
| **NIHSS** – M (SD) | 5 | 6 | 1.03 (*p* = 0.28) | 1.03 |
| **Stroke Hemisphere** (n, %)  Left  Right  Bilateral  Undetermined from Scan | 111 (37.0%)  113 (37.0%)  25 (8.0%)  53 (18.0%) | 34 (35.0%)  42 (43.0%)  7 (7.0%)  15 (15.0%) | 1.20 (*p* = 0.57)  0.92 (*p* = 0.89)  1.18 (*p* = 0.70) | 1.21  0.91  0.92 |
| **First/Recurrent Stroke** (n, %)  First Stroke  Recurrent Stroke | 209 (69.0%)  93 (31.0%) | 61 (62.0%)  37 (38.0%) | 1.01 (*p* = 0.98) | 1.36 |
| **Stroke Type** (n, %)  Ischaemic  Haemorrhagic | 248 (82.0%)  54 (18.0%) | 88 (90.0%)  10 (10.0%) | 0.50 (*p* = 0.12) | 0.52 |
| **Data-Driven Predictors** | | | | |
| **Length of Stay** – median | 8 | 10 | 1.01 (*p* = 0.61) | 1.02 |
| **Independence Prior to Admission** (n, %)  Independent  Carer Support  Family Support | 273 (90.0%)  17 (6.0%)  12 (4.0%) | 84 (86.0%)  9 (9.0%)  5 (5.0%) | 2.14 (*p* = 0.21)  0.67 (*p* = 0.61) | 1.72  1.35 |
| **Acute Mood Difficulties** (n, %)  Yes  No | 226 (75.0%)  74 (25.0%) | 77 (79.0%)  21 (21.0%) | 0.89 (*p* = 0.73) | 0.83 |
| **Charlson Comorbidity Index** – M (SD) | 1.30 (1.22) | 1.63 (1.33) | 1.08 (*p* = 0.49) | 1.22 |
| **Acute Executive Impairment** (n, %)  No Impairment  Impairment | 207 (76.0%)  64 (24.0%) | 45 (52.0%)  42 (48.0%) | **3.38 (p *<* 0.001)** | **2.78** |

**Table S16.** Descriptive statistics of model predictors between those with (*N* = 98) and without (*N* = 302) a 6-month executive function impairment.

| **Praxis Model Predictors** | **Summary Statistics** | | **Odds Ratio** | **Pooled Odds Ratio** |
| --- | --- | --- | --- | --- |
|  | **No Praxis Impairment at 6 Months** | **Praxis Impairment at 6 months** |  |  |
| **Clinically Relevant Predictors** | | | | |
| **Age at Stroke** – M (SD) | 72.81 (12.88) | 76.86 (9.96) | **1.04 (*p* < 0.01)** | **1.03** |
| **Sex** (n, %)  Male  Female | 167 (52.0%)  157(48.0%) | 50 (63.0%)  29 (37.0%) | **0.55 (*p* < 0.05)** | **0.62** |
| **Years of Education** – M (SD) | 12.32 (3.65) | 12.09 (3.33) | 0.98 (*p* = 0.56) | 0.98 |
| **NIHSS** – M (SD) | 5 | 5 | 1.00 (*p* = 0.89) | 0.99 |
| **Stroke Hemisphere** (n, %)  Left  Right  Bilateral  Undetermined from Scan | 109 (34.0%)  131 (40.0%)  31 (10.0%)  53 (16.0%) | 34 (43.0%)  27 (34.0%)  1 (1.0%)  17 (22.0%) | 0.71 (*p* = 0.31)  **0.14 (*p* = 0.06)**  0.88 (*p* = 0.75) | 0.66  **0.10**  1.03 |
| **First/Recurrent Stroke** (n, %)  First Stroke  Recurrent Stroke | 223 (69.0%)  101 (31.0%) | 48 (61.0%)  31 (39.0%) | 1.60 (*p* = 0.12) | 1.43 |
| **Stroke Type** (n, %)  Ischaemic  Haemorrhagic | 272 (84.0%)  52 (16.0%) | 67 (85.0%)  12 (15.0%) | 1.06 (*p* = 0.89) | 0.94 |
| **Length of Stay** – median | 8 | 10 | 1.00 (*p* = 0.97) | 1.01 |
| **Independence Prior to Admission** (n, %)  Independent  Carer Support  Family Support | 287 (89.0%)  27 (8.0%)  10 (3.0%) | 69 (87.0%)  4 (5.0%)  6 (8.0%) | 1.29 (*p* = 0.67)  4.66 (*p* = 0.01) | 0.62  2.49 |
| **Acute Mood Difficulties** (n, %)  Yes  No | 248 (77.0%)  74 (23.0%) | 60 (76.0%)  19 (24.0%) | 0.91 (*p* = 0.79) | 1.06 |
| **Charlson Comorbidity Index** – M (SD) | 1.37 (1.25) | 1.44 (1.33) | 1.05 (*p* = 0.66) | 1.04 |
| **Acute Praxis Impairment** (n, %)  No Impairment  Impairment | 244 (77.0%)  73 (23.0%) | 47 (61.0%)  30 (39.0%) | 1.61 (*p* = 0.13) | **2.03** |

**Table S17.** Descriptive statistics of model predictors between those with (*N* = 79) and without (*N* = 324) a 6-month praxis impairment
